# Scalable, effective, and rapid decontamination of SARS-CoV-2 contaminated N95 respirators using germicidal ultra-violet C (UVC) irradiation device

**DOI:** 10.1101/2020.10.05.20206953

**Authors:** Raveen Rathnasinghe, Robert F. Karlicek, Michael Schotsaert, Mattheos Koffas, Brigitte Arduini, Sonia Jangra, Bowen Wang, Jason L. Davis, Mohammed Alnaggar, Anthony Costa, Richard Vincent, Adolfo Garcia-Sastre, Deepak Vashishth, Priti Balchandani

**Affiliations:** Department of Microbiology, Icahn School of Medicine at Mount Sinai, New York, NY 10029, USA; Graduate School of Biomedical Sciences, Icahn School of Medicine at Mount Sinai, New York, NY 10029; Department of Medicine, Division of Infectious Diseases, Icahn School of Medicine at Mount Sinai, New York, NY 10029; The Tisch Cancer Institute, Icahn School of Medicine at Mount Sinai, New York, NY 10029; Global Health and Emerging Pathogens Institute, Icahn School of Medicine at Mount Sinai, New York, NY 10029; Smart Lighting Engineering Research Center, Rensselaer Polytechnic Institute, Troy NY 12180; Department of Electrical Computer and Systems Engineering, Rensselaer Polytechnic Institute, Troy NY 12180; Department of Chemical and Biological Engineering, Rensselaer Polytechnic Institute, Troy, NY 12180; Center for Biotechnology and Interdisciplinary Studies, Rensselaer Polytechnic Institute, Troy, NY 1218; Department of Biological Sciences, Rensselaer Polytechnic Institute, Troy, NY 12180; Department of Civil and Environmental Engineering, Rensselaer Polytechnic Institute, Troy, NY 12180; Department of Neurosurgery, Icahn School of Medicine at Mount Sinai, New York, NY, 10029; Department of Medicine, Section of General Internal Medicine, Icahn School of Medicine at Mount Sinai, New York, NY 10029; Department of Biomedical Engineering, Rensselaer Polytechnic Institute, Troy, NY 12180; Biomedical Engineering and Imaging Institute, Icahn School of Medicine at Mount Sinai, New York, NY, 10029

## Abstract

**Importance:** Particulate respirators such as N95 masks are an essential component of personal protective equipment (PPE) for front-line workers. This study describes a rapid and effective UVC irradiation system that would facilitate the safe re-use of N95 respirators and provides supporting information for deploying UVC for decontamination of SARS-CoV-2 during the COVID19 pandemic.

**Objective:** To assess the inactivation potential of the proposed UVC germicidal device as a function of time by using 3M® 8211 - N95 particulate respirators inoculated with SARS-CoV-2.

**Design:** A germicidal UVC device to deliver tailored UVC dose was developed and snippets (2.5cm^2^) of the 3M-N95 respirator were inoculated with 10^6^ plaque-forming units (PFU) of SARS-CoV-2 and were UV irradiated. Different exposure times were tested (0-164 seconds) by fixing the distance between the lamp (10 cm) and the mask while providing an exposure of at least 5.43 mWcm^-2^.

**Setting:** The current work is broadly applicable for healthcare-settings, particularly during a pandemic such as COVID-19.

**Participants:** Not applicable.

**Main Outcome(s) and Measure(s):** Primary measure of outcome was titration of infectious virus recovered from virus-inoculated respirator pieces after UVC exposure. Other measures included the method validation of the irradiation protocol, using lentiviruses (biosafety level-2 agent) and establishment of the germicidal UVC exposure protocol.

**Results:** An average of 4.38×10^3^ PFUml^-1^(SD 772.68) was recovered from untreated masks while 4.44×10^2^ PFUml^-1^(SD 203.67), 4.00×10^2^ PFUml^-1^(SD 115.47), 1.56×10^2^ PFUml^-1^(SD 76.98) and 4.44×10^1^ PFUml^-1^(SD 76.98) was recovered in exposures 2s,6s,18s and 54 seconds per side respectively. The germicidal device output and positioning was monitored and a minimum output of 5.43 mWcm^-2^ was maintained. Infectious SARS-CoV-2 was not detected by plaque assays (minimal level of detection is 67 PFUml^-1^) on N95 respirator snippets when irradiated for 120s per side or longer suggesting 3.5 log reduction in 240 seconds of irradiation.

**Conclusions and Relevance:** A scalable germicidal UVC device to deliver tailored UVC dose for rapid decontamination of SARS-CoV-2 was developed. UVC germicidal irradiation of N95 snippets inoculated with SARS-CoV-2 for 120s per side resulted in 100% (3.5 log in total) reduction of virus. These data support the reuse of N95 particle-filtrate apparatus upon irradiation with UVC and supports use of UVC-based decontamination of SARS-CoV-2 virus during the COVID19 pandemic.

## Introduction

Coronavirus disease (COVID-19) is caused by the severe-acute respiratory syndrome coronavirus 2 (SARS-CoV-2). This respiratory pathogen belonging to the beta-coronavirus group emerged at the end of 2019 in the city of Wuhan, Hubei province, China^1^. As of late September 2020, SARS-CoV-2 has infected around 31million individuals while accounting for more than 975,000 deaths worldwide^2^. So far, the United States of America is one of the hardest hit countries worldwide. The long-term public health and economic consequences are alarming and are often compared to the “Spanish-flu” pandemic caused by an influenza virus in 1918 that resulted in more deaths than there were victims in World War I^3^.

During pandemic outbreaks, frontline workers and essential workers across a spectrum of industries including but not limited to medical professionals, scientists, academics as well as individuals within food and beverage industries are required to carry out pivotal duties to keep society functioning. Critical measures that are employed to prevent the spread of the disease include avoiding social interactions through social distancing, and lockdowns in combination with improved hygiene measures. Additionally, the use of personal protective equipment (PPE) such as masks, gloves, sleeves and face-shields play an essential role in protecting front line workers as well as minimizing the spread of the respiratory pathogen SARS-CoV-2^4^. SARS-CoV-2 can spread in aerosol droplets that are formed when an infected person coughs, sneezes or even just speaks. Advanced facemasks (filtering facepiece respirators; FFR) such as N95, KN95 and N99 are used as a measure of personal protection when enhanced exposure risk to SARS-CoV-2 is expected (For instance, when working with patients or within diagnostic and research laboratories). The unexpected extent of this pandemic resulted in the long-anticipated shortage of FFR and other facemasks, the need for more PPE supplies across the world escalated significantly^5^. Such a situation negatively impacts healthcare settings, putting frontline workers at a vulnerable position, potentially resulting in a collapse of the healthcare system. This was further corroborated by the World Health Organization (WHO) stating that insufficient supply of such PPEs would put essential frontline workers at the risk of being exposed to SARS-CoV-2, which would further facilitate the spread of this contagious disease^6^. As a measure to address this issue, on the 20^th^ of March 2020, methods to conserve PPE were discussed by panelists from the journal of American medical association (JAMA) and as a result, the reuse of PPE was suggested^7^.

Though the reuse of the masks was suggested, there are several factors that one must take into consideration. Firstly, it is of importance to have a general idea in terms of the persistence of SARS-CoV-2 on surfaces. Several studies have demonstrated that viruses such as SARS-CoV and its phylogenetic-relatives such as Middle-East Respiratory Syndrome virus (MERS) and seasonal corona viruses (HCoV) may persist on inanimate object surfaces including wood^8^, plastic^9^, metal, paper and glass up to 9 days^8,9^. In contrast, a recent study suggested that SARS-CoV-2 is only able to last in such surfaces more or less up to 72 hours^10^. Taking this information into consideration, one of the solutions that was recommended was to recycle the use of respirators wherein users were recommended to use one mask per day, ensuring a grace period of five days between reuse. Given the lack of SARS-CoV-2 viral survivorship data in different surfaces, the safety and the effectiveness of the above-mentioned suggestion is largely unknown. Due to the urgency of the situation, a scalable, reliable, cost effective disinfection procedure that would enable the safe reuse of PPE is of global interest^11^.

Several methods of decontamination have been proposed in order to facilitate the reuse of PPEs. These typically branch off from chemical methods (I.e. alcohol, bleach etc.) or energetic methods such as heat inactivation, microwave-based systems or ultraviolet (UV) based systems^12^. In summary, chemical based inactivation methods have indicated that virus contaminated surfaces can be effectively decontaminated using 70% ethanol, bleach solutions (0.1% Sodium Hypochlorite) and hydrogen peroxide (0.5% H2O2) with enough exposure time^13^. The issues with such methods include inability to scale up to disinfect a large number of units and the need for specialized faculties with chemical supplies. Furthermore, several chemical treatment methods have been shown to impair the functionality of the filters found in the respirators ^14^. In contrast, energy-based systems have also been explored for decontaminating PPEs. One category of methods that was suggested was heat inactivation in which various protocols such as heating to 70°C for 24H or heating for 30 minutes at 56°C depending on the model was implemented ^15,16^. The caveat here is that this method is applicable only for a subset of respirators belonging to the FFP2 category and most manufacturers recommend against heat treatment due to its deleterious effects on the materials of the respirators which could result in ineffective protective capacity ^14,17^.

Another energy-based decontamination method is the use of germicidal UVC (200 – 280 nm) based systems^3^. UVC based disinfection methods are effective due to the fact that viral genetic materials such as RNA (relevant in this case given that SARS-CoV-2 genome is an RNA based one) and DNA strongly absorbs UVC radiation ^18,19^. This results in damage for such genetic materials at a molecular and structural level (via photodimerization), severely compromising the functionality of the virus leading to viral inactivation, consequently prohibiting the capacity for the virus to replicate upon exposure to compatible hosts^20,21^. Germicidal properties of UVC on PPE and FFRs such as N95s have been studied in the context of other respiratory viral pathogens such as influenza A viruses^22,23^. In particular, the use of UVC lamps to disinfect FFR has been studied by the National Occupational Safety and Health and found that FFR retained good filtering and air flow properties using UVC exposures that would far exceed those used to disinfect FFR indicating that UVC irradiation methods have the potential to be highly effective, point-of-use disinfection systems^24^. UVC radiation is a line of sight disinfection system and pathogens in the shadows or shielded from the UVC radiation will remain viable. The system described here is a UVC exposure system using a conveyor belt to transport contaminated FFRs between a bank of opposing UVC mercury lamps producing intense 254nm UVC radiation. The system developed here uses a vertical mask support system for UVC irradiation where both the inside and outside surfaces of the mask and straps can be dosed with UVC radiation simultaneously. The concept is shown schematically below (Fig. 1A) and a prototype has been built and tested (Fig. 1B). In the prototype, standard low-pressure Hg lamps were used as the UVC source, but UVC LED could also be used to the same effect. The system shown in figure 1B can expose contaminated N95 masks at UVC dose levels up to 1.5 J/cm2 in an exposure time of approximately one minute. The total dose can be controlled by changing the speed of the conveyor system. Dose was evaluated using commercially available color changing dosimeter tags, (Intellego Technologies, Stockholm, Sweden) which can be routinely added to the conveyor system for frequent verification of UVC dose to the exposed masks.

**Figure 1.**
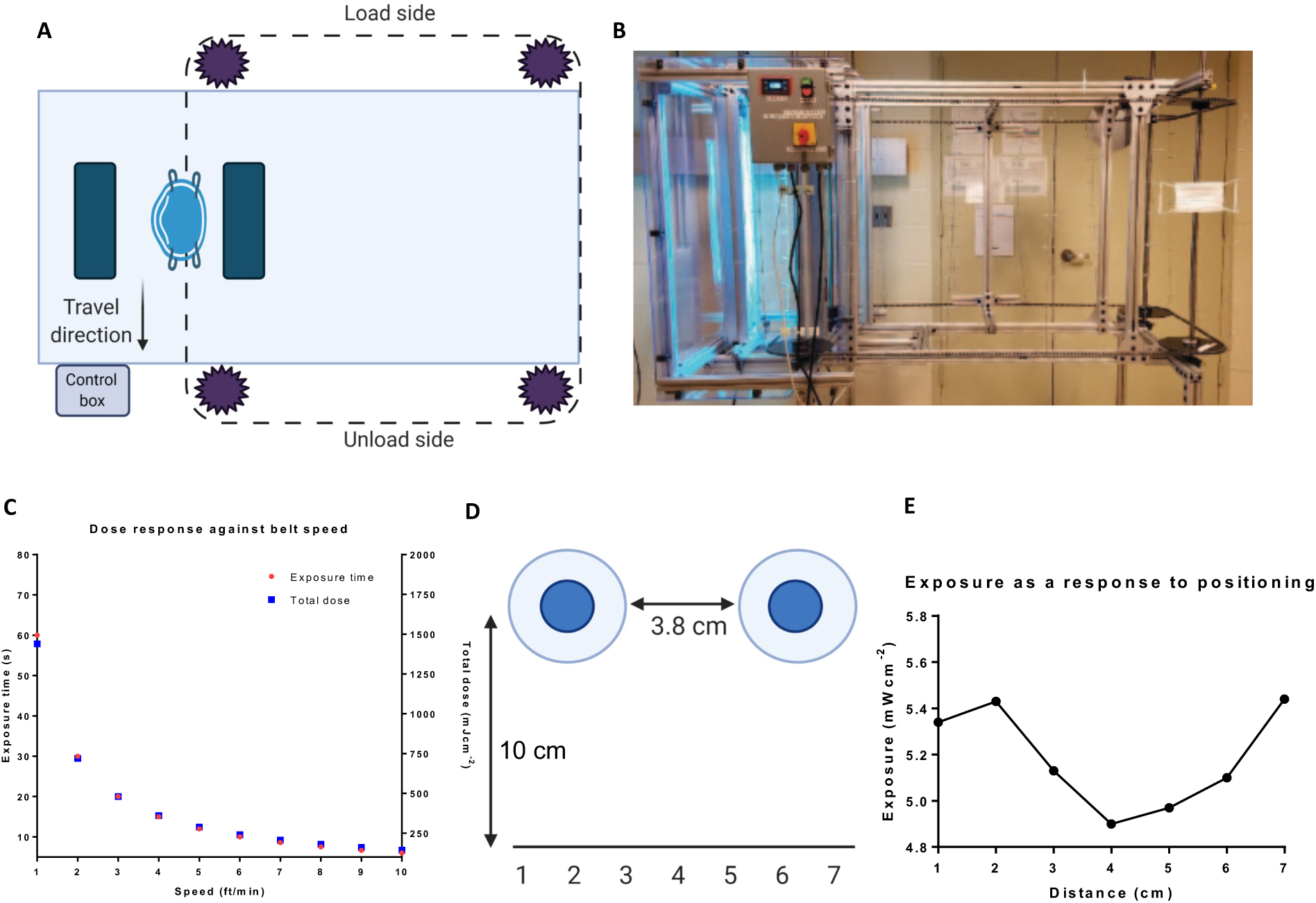
Schematic, dose response and exposure characterization of the proposed germicidal ultraviolet C (UV-C) decontamination device. **A. Schematic of the operational germicidal device**. The UVC germicidal device comprises a conveyor belt which is scalable. **B. Real image of the prototype germicidal device. C. Changes to the dose response curve as a response belt speed**. Total dose changes in response to the conveyor belt speed and exposure time. **D. Schematic of the experimental setup lamp and surface positioning**. The germicidal lamp bulbs contain 3.8 cm between them and the distance from the lamp to the surface is 10cm and exposure changes as a result of positioning was monitored. **E. Exposure profile of the lamp based on the surface placement of as indicated in D**. The optic receiver was placed 1 cm apart, covering a total distance of 7 cm and measurements were taken at 3 minutes at each time point after warming up the lamp.

Here we evaluate the effectiveness of a rapid UVC germicidal disinfection system that can be scaled up to effectively inactivate SARS-CoV-2 in potentially contaminated respirators. We demonstrate that with 240s of total exposure, an approximate reduction of 3.5 logs of SARS-CoV-2 was indicated by the lack of infective viral particles. This information provides support for use of UVC-based decontamination of SARS-CoV-2 virus during the COVID19 pandemic.

## Materials and methods

### UVC Mask Exposure System

A simple UVC mask exposure system was fabricated from two opposing IP55 high power Hg lamp fixtures (American Ultraviolet) configured vertically and a simple motorized conveyor system. The prototype system design moves FFR, supported vertically by the straps, between the two lamps where the dose is controlled by the belt speed of the mask conveyor system. The concept is shown schematically below (Fig. 1A) and a prototype was built and its mechanical operation tested (Fig. 1B).

The system shown here can expose contaminated N95 masks at UVC dose levels up to 1.5 J/cm^2^ in an exposure time of approximately one minute (Figure 1C). The total dose can be controlled by changing the speed of the conveyor system (Figure 1C). As configured the unit contains two opposing IP55 lamp fixtures, but two more pairs can be added to achieve effective irradiation at higher belt speeds for increasing FFR treatment throughput.

For dose testing with SARS-CoV-2, one lamp was removed from the system (shown in Figure 1B) and evaluated for germicidal activity in a BSL-3 facility. UVC irradiance was measured using a calibrated Optometer P 9710 Gigahertz Optik UVC meter with the 254nm detection head that was positioned at a fixed distance approximating the distance of the FFR from one of UVC lamps as configured in the UVC mask exposure system. The lamps were allowed to warm up for approximately 2 minutes to achieve stable UVC output. Due to potential masking effects that may occur in the space between the two bulbs, the changes due to positioning was assessed using 1 cm increments and experiments were done in position number 2 (Figure 1D and E).To approximate the exposure of a contaminated FFR, tests with the single lamp system were made by exposing experimental mask swatches, first on one side, then the other, as described below.

### Cells and viruses

Vero-E6 cells (ATCC® CRL-1586™, clone E6) were cultured in Dulbecco’s Modified Eagle Medium (DMEM) supplemented with 10% Fetal Bovine Serum (FBS), penicillin-streptomycin, MEM non-essential amino-acids (Gibco; 25025CL) and Hepes buffer (Gibco; 15630-080) at 37°C with 5% CO2. Cells were passaged every three days until required for experimentation. A549 human lung carcinoma cells (ATCC® CCL-185) were cultured in DMEM supplemented with 10% heat-inactivated fetal bovine serum (FBS; HyClone SH0066.03) and penicillin/streptomycin (Gibco; 15140-122) at 37°C with 5% CO2. Infectious assays using 2^nd^ generation lentivirus carrying a constitutive EGFP transgene (pLV-EGFP, Addgene) were performed in the biosafety-level 2 (BSL2) Stem Cell Core Facility at Rensselaer Polytechnic Institute. Viral stocks were produced using human embryonic kidney (HEK293) cells, with collection and concentration of LVV 48 hours after transfection of 2^nd^ generation plasmid constructs. Infectious assays for this study using SARS-CoV-2 (USA-WA1/202, bei resource – NR52281) were handled within a biosafety-level 3 (BSL3) containment facility at Icahn school of medicine at Mount Sinai by trained personnel upon approval of protocols by a biosafety committee. Propagation of viral stocks were done in Vero-E6 cell confluent monolayers by using an infection medium composed of DMEM supplemented with 2% FBS, Non-essential amino acids (NEAA), Hepes and penicillin-streptomycin at 37°C with 5% CO2 for 72 hours.

### Inactivation of N95 mask squares spiked with Lentivirus

Squares containing all layers of the 3M® 8211 - N95 particulate respirator were cut under sterile conditions resulting in final dimensions of 25mm x 25mm. N95 mask coupons were loaded with LVV-EGFP by incubation in a 50ml conical tube containing 30ml Dulbecco’s Phosphate Buffered Saline (DPBS) with 2×10^8^ transduction units (TU) of LVV-EGFP. Virus was allowed to adsorb to the mask coupons for 15 minutes. Coupons were placed in 10cm petri dishes (2 dishes, each with 4 coupons). One dish with four coupons was placed, open, under the UVC lamp with aluminum foil under the petri dish to reflect UVC light. The second dish was covered, wrapped in aluminum foil, and kept in the same biosafety cabinet without UVC exposure as a control for presence of infectious virus. The UVC lamp was placed 10 cm above the coupon surface. Coupons were exposed to UVC light for 1 minute (equivalent of 2 mJcm^-2^), turned over with sterile forceps, and exposed to UVC light for 1 additional minute. After UVC treatment, coupons were transferred to 15ml conical tubes and submerged in 5 ml of infection medium (DMEM + 1.5% heat-inactivated fetal bovine serum). Control and UVC-treated infection media were used immediately for viral infectivity assays.

### Infectivity assays

Efficiency of LVV-EGFP inactivation was tested by serial 10-fold dilutions of infection medium in the 15 ml conical tubes, followed by inoculation of 100% confluent A549 cell monolayers in duplicate. Cells were incubated for 48 hours, fixed with 4% paraformaldehyde, treated with 4,6’ diamidino-2-phenylindole (DAPI) nuclear stain for 10 minutes and imaged with a ThermoScientific Cellomics ArrayScan XTI high-content microscope to visualize infected cells as indicated by GFP. Total cell number was determined by counting DAPI+ objects and percent-infected determined by DAPI+ nuclei associated with GFP signal.

### Inactivation of N95 mask squares spiked with SARS-CoV-2

Squares containing all layers of the 3M® 8211 - N95 particulate respirator were cut under sterile conditions resulting in final dimensions of 25mm x 25mm. These were then loaded with SARS-CoV-2 by incubating them in 50 ml conical bottom tubes containing 30 ml of DMEM spiked with 1×10^6^ plaque forming units (PFU) SARS-CoV-2. The virus was allowed to adsorb into the mask coupons for 15 minutes at room temperature. Afterwards, the coupons were transferred to sterile tissue culture grade 10cm petri-dishes. These were placed over aluminum foil and under the UVC germicidal lamp after warming up the lamp. In parallel, a similar setup was conducted in which the spiked dishes were incubated in a biosafety cabinet without exposing to UV light in order to confirm the presence of virus in the N95 mask coupons. A working distance of 10cm was maintained between the lamp and the top of the coupon surface within the petri dish. Following lamp warm-up, the irradiance was measured at the working distance to be 5-8 mW/cm^2^ and was measured using the Optometer P 9710 Gigahertz Optik 254nm UVC meter and the coupons were exposed for either 2, 6, 18, 54 120, 164 and 420 seconds per side and were flipped using sterile forceps. The coupons were then transferred to 15 ml conical bottom tubes and were submerged in 3 ml of infection medium and was vortexed for one minute and were left overnight at 4°C and were assayed the following day.

### Plaque assays

Confluent monolayers of Vero-E6 cells in 12-well plate format were infected with 10-fold serially diluted samples in 1X phosphate-buffered saline (PBS) supplemented with bovine serum albumin (BSA) and penicillin-streptomycin for an hour while gently shaking the plates every 15 minutes. Afterwards, the inoculum was removed, and the cells were incubated with an overlay composed of MEM with 2% FBS and 0.05% Oxoid agar for 72 hours at 37°C with 5% CO2. The plates were subsequently fixed using 4% formaldehyde overnight and the formaldehyde was removed along with the overlay. Fixed monolayers were blocked with 5% milk in Tris-buffered saline with 0.1% tween-20 (TBS-T) for an hour. Afterwards, plates were immunostained using a monoclonal antibody cocktail against SARS-CoV-2 Spike (Creative-Biolabs; 2BCE5) and SARS-CoV-2 nucleocapsid protein NP (Creative-Biolabs; NP1C7C7) at a dilution of 1:1000 followed by 1:5000 anti-mouse IgG monoclonal antibody and was developed using KPL TrueBlue peroxidase substrate for 10 minutes (Seracare; 5510-0030). After washing the plates with distilled water, the number of plaques were counted. Data shown here is derived from three independent experimental setups.

## Results

### Inactivation of N95 mask squares spiked with LV-EGFP

Supernatants derived from untreated mask snippets were used to infect A549 cells and a total of 36001 cells were analyzed. A total of 2.57% cells (approximately 925 cells) were GFP positive because of LV-EGFP infection and constituted around 1155 viral transduction units per ml of analyzed supernatant (Figure 2A). Supernatants derived from snippets treated for 60 seconds each side showed only around 0.08% GFP positive cells (approximately 27 cells out of a total of 33908) accounting for 34 viral transductions per ml (Figure 2B).

**Figure 2.**
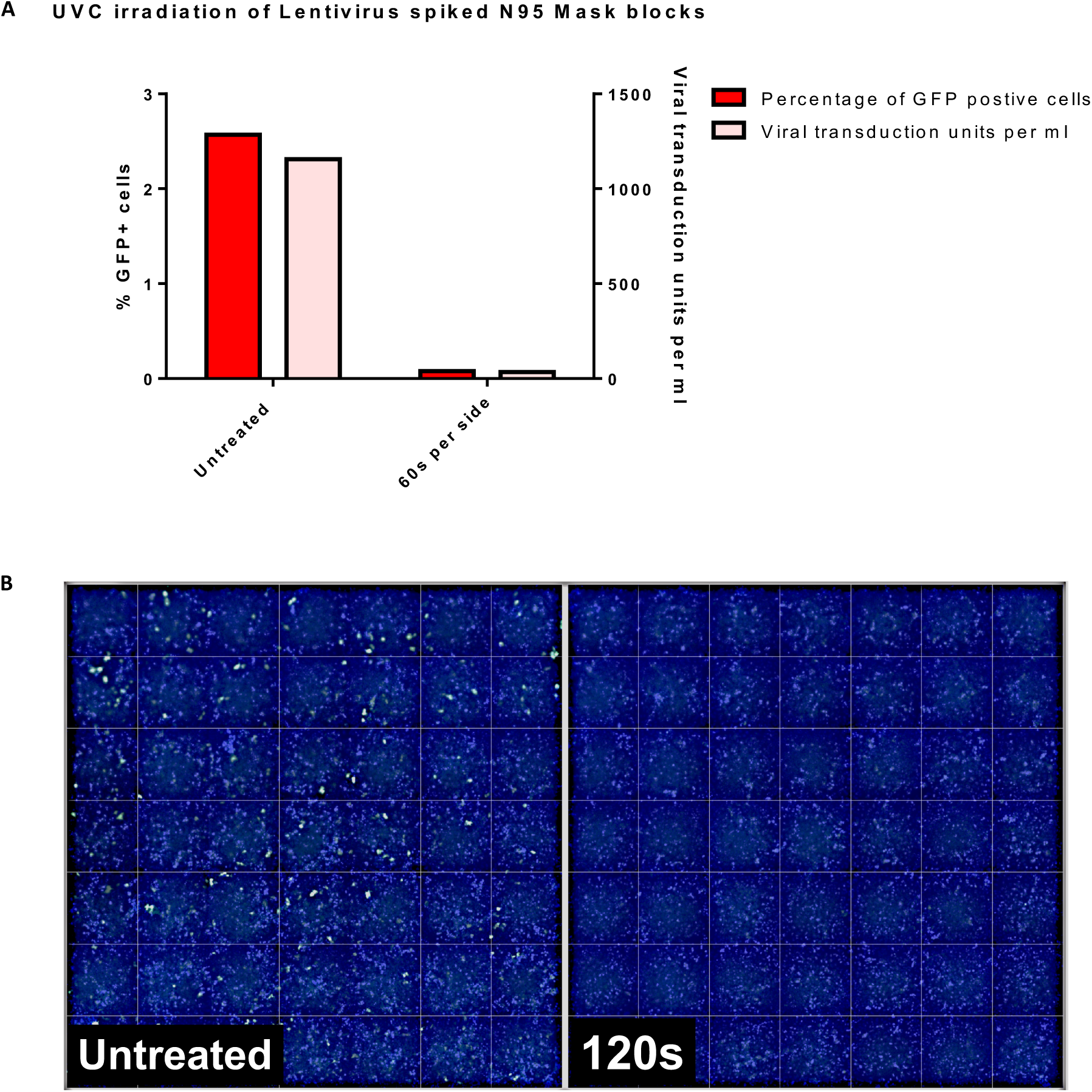
Ultraviolet C (UV-C) exposure of N95 mask squares spiked with LV-EGFP indicated inactivation of lentivirus after treatment for a total of seconds. N95 mask squares (2.5cm^2^) with all layers were incubated with infection media spiked LV-EGFP for 15 mins. The masks were then placed on open 10cm polystyrene cell culture dishes 10 cm from the lamp. After warming up the lamp, the blocks were treated for 60 seconds per each side. Masks were left in infection media at 4°C overnight. 1 ml of the mask derived media mix was used from each condition and used to infect A549 cells, cells were fixed with 4% formaldehyde and was stained with DAPI and was imaged using a Cellomics ArrayScan. **A. UVC treatment of N95 snippets spiked with LV-EGFP indicates viral inactivation after 120s of total exposure**. GFP+ cells and the total viral transduction units per ml of supernatant was assessed and data shown here is derived from two independent experiments. **B. Fluorescence microscopy analysis of infected A549 cells indicate viral inactivation**. Supernatants derived from treated or untreated 3M-N95 snippets were used to infect A549 monolayers. 48HPI, the cells were fixed and were stained for nucleus using DAPI (blue)

### Total inactivation of SARS-CoV-2 in spiked N95 apparatuses by 120 seconds of exposure per side

The warmed up UVC device was able to neutralize approximately 1 log viral titer with as little as 2 seconds of exposure per side (Figure 2A). Further reduction of approximately 2 logs was observed by an exposure of 54 seconds and by 120 seconds (per side). Total inactivation was apparent by the lack of plaque forming units (PFU) derived from the respective experimental settings. This was achieved by using an irradiance of at least 5.43 mW/cm^2^. Figure 3 demonstrates exponential decay (r^2^=99; p<0.001) in PFUs and fraction of active virus remaining as function of both exposure times and dose. While we observed a significant reduction by 54 seconds of exposure per-side, we were unable to replicate it in one of the three independent experiments.

**Figure 3.**
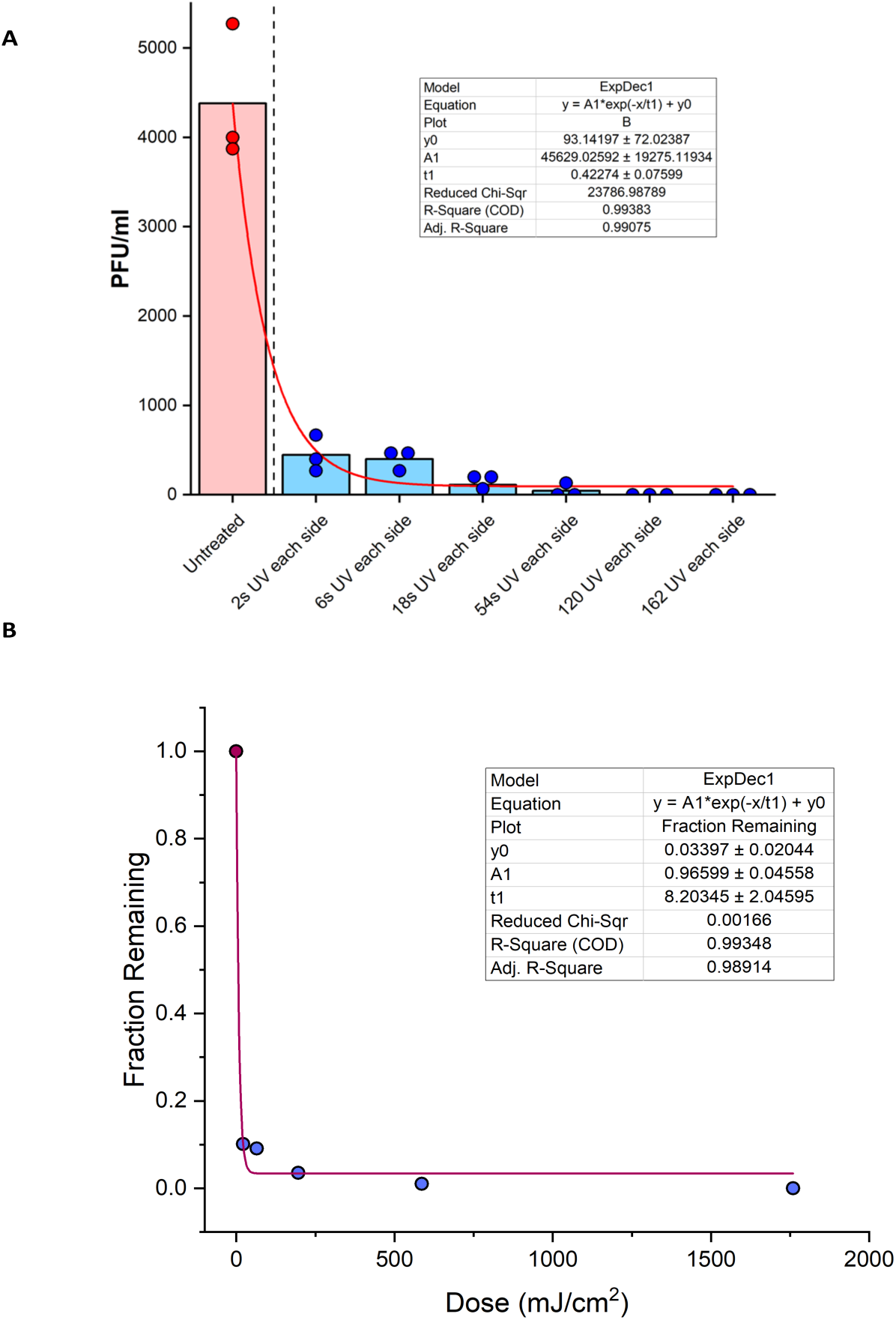
UVC irradiation for 240 seconds inactivates over 3.5 log reduction in SARS-CoV-2 viral titers. **A. Time course log reduction of SARS-CoV-2 viral reductions of N95 snippets treated with UVC irradiation**. N95 mask squares (2.5cm2) with all layers were incubated with infection media spiked SARS-CoV-2 for 15 mins. The masks were then placed on open 10cm polystyrene cell culture dishes six inches from the lamp. After warming up the lamp, the blocks were treated for the indicated amount of time per each side. Masks were left in infection media at 4°C overnight. 1 ml of the mask derived media mix was used from each condition and was titered VERO-E6 cells using standard plaque assays and were developed mAb cocktail composed of SARS-CoV-2 Spike (S) and Nucleocapsid protein (NP). Each dot represents data from three independent experiments and plotted data bars represents mean and standard deviation for the triplicates. Dotted line is the minimal level of detection (approximately 67 PFUml-1). Baseline for the assay is shown using a continuous line. **B. Regression analysis of viral reduction as per timely UVC irradiation**.

## Discussion

COVID-19 pandemic caused by SARS-CoV-2 has negatively impacted the entire world, affecting all industries across multiple disciplines. All essential workers, particularly healthcare workers, extensively rely on FFRs such as N95 respirators in order to minimize the risk of contraction and transmission of SARS-CoV-2. Given the rapidly expanding nature and this biologically hazardous agent, a high use and demand for such respirators is evident^25^. Owing to those reasons, such respirators are at the risk of running into a considerable shortage, suggesting the need for safe reuse. Based on the limited number of SARS-CoV-2 virus survivorship data coupled with the need to preserve FFRs, the government of the USA recommended the use of one respirator per day, cycling them throughout the week ensuring a minimum of five days between reuse^26^. The risk associated with this due to the largely unknown virus survivorship data urges the need for not only more survivorship studies, but also studies that would explore the effective decontamination of facepieces. This has encouraged researchers to evaluate several potential methods for the decontamination of FFRs. Thus, chemical-based methods including the use of alcohol, bleach, ethylene oxide, ozone decontamination and H2O2 based methods have developed to effectively decontaminate respirators^12,14,17,18,22^. On the other hand, the use of energy-based systems such as dry heat, microwave, gamma-radiation and ultraviolet based germicidal systems have also been tested. However, there are two important factors to consider in such instances; 1) any disinfection method used should guarantee the complete inactivation of SARS-CoV-2 and 2) treatment of choice should not have deleterious effects that may impair the functionality of the respirator^7^. Consequently, scientists, manufacturers and professionals in the medical field are working towards devising a strategy that would address the aforementioned factors.

Amongst the methods of decontamination, ultraviolet C germicidal irradiation (UVGI) has a lengthy history of antimicrobial applications in healthcare and medical based settings^27^. Due to its scalable ease, UVC based decontamination methods are favorable during a pandemic setting such as the current SARS-CoV-2 pandemic and can be deployed more generally, for example to decontaminate public spaces. Here, we evaluated a germicidal UVC irradiation system as a potential tool to decontaminate respirators with the aim of assisting the safe reuse of them.

We first characterized the device by obtaining the dose response parameters as well as identifying the importance of the positioning of the masks which are to be sterilized in order to obtain the most efficient decontamination setting (Figure 1C, D and E).The prototype derivative we had was first used to test the germicidal potential, using a lipid enveloped 2^nd^ generation Lentivirus as a model virus under biosafety level 2 containment. The data suggested robust reduction of lentivirus from the supernatants that were infused with the N95 snippets upon UVC treatment for a total 120 seconds (Figure 2). By taking the above data into consideration, we then evaluated a time and dose dependent UVC irradiation protocol for the decontamination of N95 mask snippets spiked with live SARS-CoV-2 under BSL3 conditions. Of note, two seconds per side of irradiation (4 seconds total exposure time) indicated the reduction of approximately 1 log of viral titer with a minimal irradiance of 5.43 mW/cm^2^. While we were able to see an approximate 2.5 log reduction of mask snippets treated for a total of 108 seconds (54 s per side) in two out of three independent experiments, we were able to see the reduction of approximately 3.5 log viral reduction by 240 seconds of total UVC irradiation in all experimental replicates. An important note to highlight here is that we used a functional assay such as plaque assay to test the inactivation as opposed to utilizing a technique such as qPCR. Molecular techniques as such may not truly reflect the degree of inactivation given that the presence of nucleic acids such as RNA is not indicative of live virus.

Although we spiked the snippets with 10^6^ PFU of SARS-CoV-2, we were only able to retrieve an average viral titer of 4.3×10^3^ PFU. This can be accounted to the fact that the spiked snippets were suspended in a larger volume of 15 ml after spiking. Our study was able to demonstrate a 3.5 viral log reduction by 240s of irradiation, yet it is currently unknown as to the amount of viruses that may be present in FFRs derived from real-life settings such as clinics and high biosafety level laboratories. Albeit the promising data in the current study, several limitations warrants further investigation in this matter. For instance, N95 manufacturers such as 3M do not recommend the decontamination as well as the reuse of FFRs. This probably has been suggested based on a study conducted by 3M in which respirators were exposed to cumulative dose of 10 Jcm^-2^ between five to 10 cycles resulting in changes in mask fit and physical characteristics^28^. This is indicative of the fact that FFRs are only able to withstand a certain amount of decontamination cycles before the performance is compromised. Furthermore, studies have suggested that repetitive donning and removal FFRs result in impaired mask fit^29^, indicating that such respirators can only be reused a very small amount of time regardless of the decontamination efficiency. While there are no strict guides explaining the possible number of reuses for FFRs, the CDC recommends no more than 5 reuses based on two studies ^29-31^. Therefore, a non-deleterious germicidal protocol can be implemented for the decontamination of FFRs to support the safe reuse for a limited number of times. Other studies have demonstrated the effects on the materials and fit upon UV-C irradiation using doses 1-1.2 Jm^-2^. It was noted that there was no significant damage to the respirators, compromising the performance. However, some impact on fit due to strap degradation was noted after an unspecified number of cycles and was not quantitatively addressed 14,17,19,24. Furthermore, a variety of N95 FFRs in the range considered here (1-1.2 J/cm^2^) were tested for performance post treatment and found no significant respiratory filter material damage, impact on filtration performance. The proposed device in the current study will be operating below these dosages and exposure ranges.

The current COVID-19 pandemic has presented a multitude of challenges in healthcare settings and the seemingly apparent shortage of the PPE such as FFRs is currently an issue as such. This study evaluated UV-C based germicidal decontamination protocol which has the potential to inactivate 3.5 log SARS-CoV-2 within a total time frame of 240s, facilitating a potential avenue for the safe reuse of the FFRs.

## Data Availability

Data can be made available through email request to the corresponding author, Dr. Priti Balchandani.

## Acknowledgments

We thank Randy Albrecht for support with the BSL3 facility and procedures at the ISMMS, and Richard Cadagan for technical assistance. This research was partly funded by CRIP (Center for Research for Influenza Pathogenesis), a NIAID supported Center of Excellence for Influenza Research and Surveillance (CEIRS, contract # HHSN272201400008C); by the generous support of the JPB Foundation, the Open Philanthropy Project (research grant 2020-215611 (5384)), anonymous donors to AG-S, funds from Rensselaer Polytechnic Institute and NIH (5UL1TR001433).

